# Evaluation of the Point of Care Molecular Diagnostic Genedrive HCV ID Kit for the detection of HCV RNA in clinical samples

**DOI:** 10.1101/2020.05.10.20097576

**Authors:** Abhishek Padhi, Ekta Gupta, Gaurav Singh, Reshu Agarwal, Manoj Kumar Sharma, Shiv Kumar Sarin

**Affiliations:** Department of Clinical Virology, ILBS, New Delhi, India. 110070; Department of Hepatology, ILBS, New Delhi, India. 110070

**Keywords:** Hepatitis C, Point of care test, HCV care cascade

## Abstract

2.

**BACKGROUND:** Despite the availability of an effective treatment for HCV infection, major bottle neck in the HCV care cascade is difficulty in getting the HCV RNA done on serologically positive (antibody to HCV) patients. This is due to the limited availability of HCV RNA test in the periphery. Nucleic acid amplification testing at point of care might revolutionise the HCV care continuum through its increased sensitivity, decreased turn around time and ease of performance in the field. Genedrive is one such assay which has recently got a CE- IVD certification.

**OBJECTIVES:** The diagnostic accuracy of Genedrive HCV ID kit for the qualitative detection of HCV RNA was evaluated by comparing with Abbott HCV RNA in an Indian demographic setting and across a range of different genotypes commonly found in India.

**STUDY DESIGN:** For the assay evaluation 150 HCV RNA positive and 170 HCV RNA negative samples as tested by Abbott HCV RNA assay (Abbott, Wiesbaden, Germany) on Automated m2000sp/m2000rt platform, as per the manufacturer’s instructions, that were retrieved from −80°C in the Virology repository were included in the study. All the samples were retrieved and re tested for HCV RNA on Genedrive HCV ID kit and compared with Abbott HCV RNA results.

**RESULTS:** Comparison of the Genedrive HCV ID kit with the Abbott Real Time HCV assay revealed a sensitivity of 100 % (95% CI 97.9 to 100) and a specificity of 100 % (95% CI 97.9 to 100) with a positive predictive value and negative predictive value of 100%. Overall diagnostic accuracy of Genedrive was found to be 100% (95% CI 98.9 to 100).

**CONCLUSION:** This study demonstrates that Genedrive HCV ID kit can be used for decentralised testing of HCV and ultimately finding the missing million.

**Highlights:**

- Decentralisation of HCV RNA assays is the need of hour for finding the missing millions.
- Genedrive HCV ID kit is one such assay which can qualitatively detect HCV RNA and can be used as a point of care molecular assay thereby reducing the dropouts and improving the HCV care cascade.

## 3. Background

Hepatitis C virus (HCV) infection known for its chronicity resulted in approximately 3,99 000 death in 2016 due to liver cirrhosis and hepatocellular carcinoma [1]. Globally an estimated 71 million individuals are infected with HCV out of which India is one of the six countries (others being China, Pakistan, Nigeria, Egypt & Russia) where more than 50% of the infected population resides[2]. A recent meta-analysis of HCV infection in Indian population predicted around 3 – 9 million persons with active Hepatitis C infeetion[3].

In 2016, the world health organization (WHO) launched its ambitious strategy to eliminate viral hepatis as a public threat by 2030 with a targets of 90% reduction in incident cases of hepatitis B and C and a 65% reduction in mortality[4]. In absence of vaccine against HCV and with the introduction of directly acting antivirals (DAA) as an effective treatment option, the onus squarely lies on an accurate diagnostic modality to achieve this target.

The traditional approach to HCV diagnosis requires an HCV antibody test followed by detection of HCV RNA to confirm viremia in HCV antibody (HCV-ab) positive cases[1]. However, HCV-Ab detection includes individuals who have cleared infection either spontaneously or through treatment (estimating exposure as well as active infection), and also includes false positives [5]. Furthermore low signal upon cut off presents a diagnostic dilemma in the detection of HCV-ab[6]. Again the rapid diagnostic tests pitted as the point of care test (POCT) have varied sensitivity in different settings [7] [8]. As a result, there has been a paradigm shift towards using HCV PCR to determine accurately the population prevalence of active infection [5].

While HCV RNA testing allows determining current status of infection thus indicating individuals for treatment[9], it is confined to few high-end laboratories. This centralized arrangement of HCV RNA testing may result in patients losing to the follow up. Presently, the Cepheid Xpert HCV Viral Load assay (Cepheid) is currently the only CE-IVD certified assay for decentralized HCV VL determination. Though the Xpert HCV reported to have a good performance, a high sample volume and need of electrical power supply limits its field use[10]. Hence a robust, fully automated, high throughput platform with a low turn around time is the need of the hour that can serve as a single point molecular testing and refer to care centre so that loss to follow up can be prevented particularly in the grass root level.

The Genedrive HCV ID Kit is one such platform which is European Union CE approved, hand held battery operated fully automated platform developed as a point-of-need molecular test for confirmation of chronic HCV. The test is a cartridge based reverse transcriptase polymerase chain reaction (RT-PCR) with an end point melt analysis of fluorescent molecular probes. It has so far been validated in European and African settings with the predominant HCV genotype as genotype 1,3,4 and 5, and has demonstrated good diagnostic accuracy (sensitivity 99.8 96, specificity 100%) [10]. The Genedrive HCV ID Kit requires validation in Indian context where the circulation of HCV genotype is predominately 3 and 1 to ensure suitability for the Indian population.

## 4. Objectives

The objectives of this study were to determine the diagnostic accuracy of the Genedrive HCV ID Kit for HCV RNA detection, as a confirmatory test for seropositive HCV patients in Indian demographic settings.

## 5. Study Design

### 5.1. Samples

Samples were retrieved from −80 from the Virology data base, 150 positive and 170 as negative samples. All reference test data were collected retrospectively, whilst Genedrive data were collected prospeetively. All specimens were remnants from routine testing of participants either coming to the outpatient department or admitted in the hospital. The specimens were plasma derived from blood collected in EDTA vacutainers that had been stored at −80°C and were collected at ILBS between date (October 2017) and date (Feb 2019).

Cases were determined as serologically positive for HCV with confirmation of HCV RNA presence by the Abbott Real Time HCV assay (Reference Test, henceforth called Abbot assay). Whilst the controls were determined as serologically positive for HCV with confirmation of HCV RNA absence by the Abbott assay (Reference Test).

HBV and HIV co-infected samples and samples with inadequate volume were excluded from the analysis.

### 5.2. Abbott Real Time HCV assay

The Abbott assay was considered as reference test for detection of HCV RNA in plasma samples. The test is quantitative with a 95% limit of detection (LOD) of 12 IU/mL and measuring range of 12 to 10^8^ IU/mL. 750 μL of plasma per specimen was analysed using the Abbott assay on the automated m2000sp/m2000rt platform, as per the manufacturer’s instructions.

### 5.3. Genedrive HCV ID Kit

In the Genedrive assay 30 of plasma sample was added to the cartridge as per the manufacturer’s instructions. The Genedrive assay has a lower limit of Detection of 3.37 log_10_ IU/mL (2362 IU/mL) and kit has been validated to work on all the genotypes(as per the kit literature). [11].

### 5.4. HCV Genotyping

Wherever a viral load of >3 log_10_ value was their the genotyping was done. Genotyping of positive HCV RNA samples was performed as described previously [12]. Briefly, genotyping of HCV RNA positive samples was performed using the 5’UTR region of the viral RNA. A farther confirmation of genotype 1 samples was performed using the NS5B RNA region [13]. Genotyping of amplified and purified DNA was performed by bidirectional sequencing using ABI Big Dye chemistry on the ABI 3500DX series genetic analyzer (Life Technologies, Waltham, MA). Sequence reads were aligned using DNA Baser V3.5.1 (Heracle BioSoft SRL, Romania). A Basic Local Alignment Search Tool (BLAST) was performed using obtained sequences using the database of NCBI.

### 5.5. Statistical analysis

Statistical analysis was done using the Statistical SPSS software, version 22.0; Chicago, IL, USA). Continuous variables were expressed as mean ± SD or median (range) as appropriate and categorical variables were expressed as percentage. All statistical tests were 2-tailed, and results were statistically significant with a p value was less than 0.05.

#### Ethics

The study was approved by the Institutional ethics committee (IEC) of Institute of Liver and Biliary Sciences (ILBS) and patient consent form was waived off because of the use of left over clinical samples sent to virology laboratory for routine diagnosis for HCV.

## 6. Results

### 6.1. Sample details

A total of 320 samples were included in the study, the baselines characteristics of the study population is described. Of the 150 HCV positive specimens, thirteen were below the Genedrive 95 % Limit of Detection of 3.37 log_10_ IU/mL (2362 IU/mL), whilst the remaining 137 were above. Genotyping could be done in 105 samples.

**Table 1.**
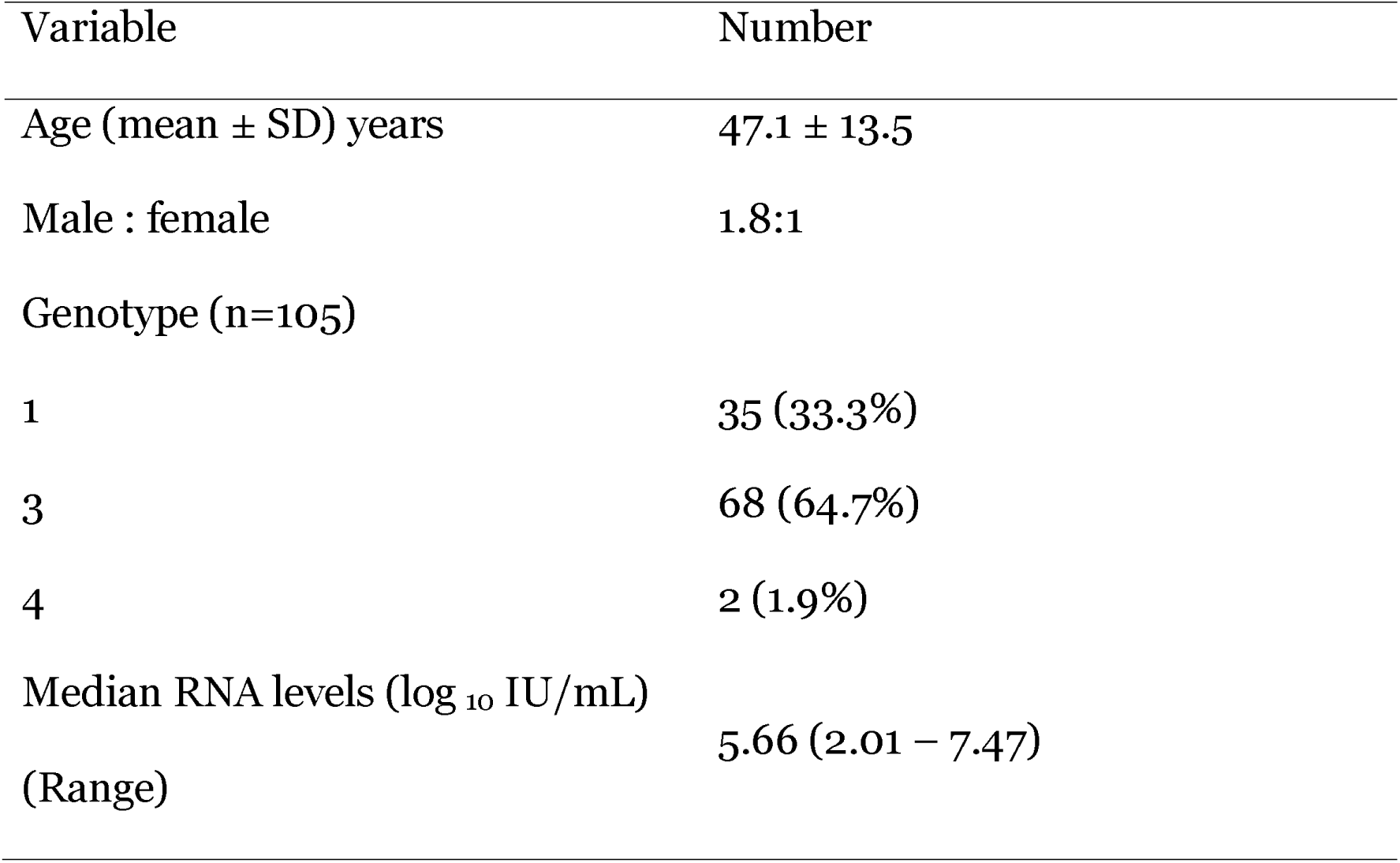
– Baseline characteristics of the study population

### 6.2. Diagnostic accuracy

Comparison of the Genedrive HCV ID Kit with the Abbott Real Time HCV assay revealed a sensitivity of 100% (95% CI 97.6 to 100.0) and a specificity of 100% (95 % CI 97.9 to 100), with a Positive Predictive Value and an Negative Predictive Value of 100%. The overall diagnostic accuracy of the Genedrive was determined at 100% (95% CI 98.9 to 100.0). Thirteen specimens were included in the study below the stated Genedrive 95% Limit of Detection of 3.37 log_10_ IU/mL, these specimens had a median viral load of 2.96 log_10_ IU/mL and a range of 2.01 − 3.32 log_10_, IU/mL. All thirteen specimens were detected by Genedrive.

The lowest viral load that was detected by genedrive was 103 IU/ml (2.01 log_10_IU/ml).

**Table 2.** – Comparison of HCV RNA detection between Abbott and Genedrive assay.

| Abbott |  |  |  |
| --- | --- | --- | --- |
| Undetecte | $<3.37 \log_{10}$ | $>3.37 \log_{10}$ | Total |

|  |  | d | IU/mL | IU/mL |  |
| --- | --- | --- | --- | --- | --- |
|  | Undetectd | 170 | 0 | 0 | 170 |
| Genedrive | Detected | 0 | 13 | 137 | 150 |
|  | Total | 170 | 13 | 137 | 320 |

### 6.3. Discrepant samples

No discrepancy was observed between Genedrive with the Abbott HCV assay.

## 7. Discussion

The present study aimed at evaluating the diagnostic accuracy of Genedrive HCV assay for the qualitative detection of HCV RNA. A comparison of Genedrive was done with Abbott Real Time HCV test, which offers viral load and genotype testing and is currently considered as the gold standard. Sensitivity of 100% (95% CI 97.6 to 100.0) and a specificity of 100% (95% CI 97.9 to 100) was observed in our study. Studies done by Libre *et al* [10] found high diagnostic sensitivity of 98.6% (95% CI 96.9% to 99.5%) and 100 *%* specificity (95% CI 99.3% to 100%) to detect HCV viraemia in Western and South African laboratories.

WHO recommends that HCV serology testing be offered to individuals who are part of a population with high HCV prevalence or who have a history of HCV risk exposure/ behaviour and confirm the diagnosis of chronic HCV infection by nucleic acid testing (NAT) for the detection of HCV RNA [9]. Currently HCV RNA testing is available in only few of a centralised medical set ups resulting in < 1% of infected individual being aware of their infection [14]. In Europe only 4.1% of individuals have been cured from initial viraemia [15] and the situation is even more grim in resource limited countries where it is <1 %[16][17]. In countries like Egypt and Mongolia the prevalence of HCV is very high but the treatment rate is appalling, 0.12% and 1.2% respectively, thereby suggesting that there is a negative correlation between HCV prevalence and treatment[18][19]. Hence there is an urgency in the need of a point of care molecular test which can be an alternative to the conventional HCV NAATs, that can be used in the field thereby decentralising the HCV diagnosis mainly in the resource limited countries.

The Genedrive is a portable, fully integrated, PCR-based platform. Genedrive has been designed to enable ‘Direct to PCR’ analysis of sample matrices without the requirement to undertake pre-extraction of DNA, all in approximately 90 minutes from sample collection. Genedrive is a 600 gram handheld thermocycler that qualitatively detects HCV RNA by NAAT in plasma and serum samples. It fulfils the target product profile of HCV diagnosis drafted by Foundation for innovative new diagnostics (FIND)/WHO (specificity > 98% and sensitivity > 95%) and has shown diagnostic accuracy for all 6 major genotypes. The limit of detection (LoD) of Genedrive HCV RNA assay is 2362 IU/ml as compared to Abbott HCV assay which is 12 IU/ml. But, recent WHO guidelines suggest that “a limit of detection of 3000 IU/mL or lower would be acceptable and would identify 95% of those with viraemic infection”[20]. The Genedrive HCV ID Kit can potentially contribute to a decentralisation of clinical management of chronic hepatitis C, which may result in expansion of the treatment programme to rural areas of resource limited settings due it its relative ease of use in comparison to current centralised laboratory based systems for detection of HCV RNA.

Currently there are several NAAT point of care testings in pipeline and the Cepheid Xpert HCV viral Load assay is currently the only CE- IVD certified assay for decentralised HCV viral load determination. Despite having good performance Xpert HCV test presents important limitations. The Xpert HCV assay requires large volume of sample (1 ml), a constant power supply, for which a basic laboratory set up is required. Moreover, the catridge of Xpert HCV assay requires guanidium thiocyanate as lysis reagent which is highly toxic and requires special care while handling and disposal [12] [21].

In contrast Genedrive is a battery operated handheld machine requiring minimal amount of sample (30μl) and there is no toxic chemicals which makes Genedrive ideal to be used in field settings.

The present study was carried out in a tertiary care centre and the test assay was performed by highly trained technicians in a controlled environment. Furthermore performance of Genedrive HCV assay requires a prospective validation in a real life decentralised peripheral resource limited setting in low- and middle income countries.

Finally, an important factor when assessing the feasibility of Point of care technologies is their cost, both for platform and assays. However, direct comparisons of costs to other platforms are challenging as there are geographic differences and unknowns like subsidised or negotiated pricing, distributor margins and duty. Furthermore, additional costs associated with the platform should be considered, including installation, maintenance, training and lifespan of the equipment and tests. While the ultimate Genedrive costs remain to be defined, the instrument is approximately $5000, which is considerably less than other molecular systems, and every Genedrive HCV test are going to be available for $30–$40, counting on the country or region in question.

## 8. Conclusions

Despite the lack of quantification of HCV viral load Genedrive HCV assay offers a great potential to be used as a point of care NAAT thereby enabling real time diagnosis, minimising loss to follow up and prompt management of patients with chronic HCV infection in any clinical setting.

## CRediT authorship contribution statement

**Abhishek Padhi**: Writing - original draft, Writing - review & editing, Methodology. **Ekta Gupta** - Conceptualization, Formal analysis, Investigation, Data curation, Writing - original draft, Writing - review & editing, Supervision, Project administration. **Gaurav Singh**: Resources, Methodology. **Reshu Agarwal**: Data curation. **M Sharma**: Writing - review & editing, Supervision. **Shiv Kumar Sarin**: Project administration, Supervision, Writing - review & editing.

## Data Availability

No external data sets.

## Acknowledgement

We thank Genedrive plc for supply of the instrument and Genedrive® HCV ID Kit.

